# Identification of Three Endotypes in Pediatric Acute Respiratory Distress Syndrome by Nasal Transcriptomic Profiling

**DOI:** 10.1101/2020.04.28.20083451

**Authors:** James Garrett Williams, Rashika Joshi, Rhonda L. Jones, Aditi Paranjpe, Mario Pujato, Krishna Roskin, Toni M. Yunger, Erin M. Stoneman, Patrick M. Lahni, Hector Wong, Brian M. Varisco

## Abstract

Acute respiratory distress syndrome (ARDS) and pediatric ARDS (PARDS) can be triggered by multiple pulmonary and non-pulmonary insults and are the source of substantial morbidity and mortality. The nasal and lower conducting airways have similar cell composition and nasal transcriptomes identify disease state and sub-classes in lung cancer, COPD, and asthma. We conducted an observational, prospective trial to determine whether this technique could identify PARDS endotypes in 26 control and 25 PARDS subjects <18 admitted to the pediatric ICU. RNA from inferior turbinate brushing was collected on days 1, 3, 7, and 14. Standard RNA-processing yielded 29% usable specimens by mRNA-Seq, and a low-input protocol increased yield to 95% usable specimens. 64 low-input specimens from 10 control and 15 PARDS subjects were used for model development. Control and some PARDS subjects clustered together in Group A while some day 1, 3, and 7 specimens clustered into Groups B and C with specimens from these subjects moving to Group A with PARDS resolution. In multivariate analysis, the only clinical variables associated with specimen Group B or C assignment was severity of lung injury or viral PARDS trigger. Compared to Group A, Group B had upregulation of innate immune processes and Group C had upregulation of ciliary and microtuble processes. Analysis of the 15 standard processing specimens identified the same grouping. Mortality trended higher in group B (25%) and C subjects (28.6%) compared to A (5%, p=0.1). Comparison of groups with 16 PARDS-associated serum biomarkers identified correlation of Endotype B with Tumor Necrosis Factor-*α*, but not other inflammatory cytokines and Endotype C with Surfactant Protein D. We identified three nasal transcriptomic PARDS endotypes. A is similar to control. B is marked by an innate immune signature only weakly reflected in the serum. C may be associated with loss of epithelial barrier integrity. Nasal transcriptomics may be useful for prognostic and predictive enrichment in future PARDS trials. ClinicalTrials.gov Identifier NCT03539783

## 3 Background

In the Pediatric intensive care unity (PICU), pediatric acute respiratory distress syndrome (PARDS) is a leading source of morbidity and mortality ^1^. Despite decades of research and many large, multicenter, randomized clinical trials in ARDS, the only consensus therapies are supportive: the use of low tidal volume ventilation and employing a restrictive fluid strategy ^2 3^. A criticism of many studies in ARDS has been their failure to account for etiologic and physiologic differences ^4^. ARDS etiology can be broadly categorized as direct (i.e. pneumonia, aspiration, etc.) or indirect (i.e. sepsis, hemorrhagic shock, etc.) In adult ARDS, both direct and indirect ARDS confer similar mortality despite lower illness severity scores in direct ARDS, suggesting different pathophysiological processes ^5^; differences that are dramatically exemplified in a recent publication showing near-complete replacement of epithelial cells with macrophages upon lung biopsy in two young adults placed on extracorporeal membranous oxygenation (ECMO) for presumed community acquired pneumonia ^6^. However, categorizing the inciting injury as direct or indirect has proven inadequate to guide therapy ^7 8^. Nonetheless, several serum biomarkers suggest differences in the pathophysiology of direct vs. indirect ARDS ^9^ suggesting their potential use to identify PARDS endotypes. Elevated serum levels of Receptor for Advanced Glycation End-product (RAGE, also termed AGER) has been reported to predict ARDS progression ^10^, and the endothelial cell protein angiopoietin-2 (ANG2) was positively associated with mortality in PARDS ^11^ and adult surgery or trauma-related ARDS ^12^. Elevated serum Matrix Metalloproteinase (MMP) level have been associated with PARDS mortality ^13^. In latent group analysis of a clinical trial of fluid administration in ARDS (FACTT trial) ^3^, subjects with higher levels of inflammation (as evidenced by higher interleukin-8 (IL8) and tumor necrosis factor receptor-1 (TNFRSF1) and lower serum bicarbonate levels, were less likely to survive with a fluid liberal strategy, while subjects with lower levels of inflammation were more likely to survive ^14^. Hyperinflammatory ARDS patients were also found to have worse outcomes with higher positive end expiatory pressure (PEEP) levels ^15^. Thus, serum biomarkers demonstrate the ability to differentiate two ARDS sub-types, could help to guide therapy, and predicted higher mortality.

However, there is perhaps a limit to the extent to which peripheral blood-based assays accurately reflect lung pathology. While peripheral blood gene expression profiling in pediatric sepsis had identified important endotypes that correlate with outcome ^16^, there is only ~40% concordance of leukocyte gene expression between lung and peripheral leukocytes with ~20% of genes being expressed discordantly ^17^. While RAGE is a lung-specific protein, ANG2 and inflammatory cytokines are present in multiple tissues and may lack specificity. We propose that directly assaying the gene expression of respiratory epithelial cells can better characterize lung pathobiology and longitudinally assessing this expression to identify key pathways and processes in PARDS recovery. Such profiling has been well-established in other respiratory diseases. Bronchial and nasal gene expression profiling was highly diagnostic for the presence of lung cancer in smokers ^18^, and for corticosteroid sensitivity in asthma ^19^. Nasal and bronchial gene expression profiling can differentiate between COPD and non-COPD in smokers ^20^. To accomplish this in the PICU environment, collection, storage, transportation, and processing techniques must minimize the degradation of sample RNA and be compatible with the ICU environment. Here, we report the development of a technique to collect quality RNA specimens from nasal respiratory epithelial cells and present data that nasal epithelial cell transcriptomics can be used to identify three distinct PARDS endotypes.

## 4 METHODS AND DESIGN

Comprehensive methods can be found in the online supplement.

### Human Subjects Research

Studies were approved by the Cincinnati Children’s Hospital Institutional Review Board (IRB 2015-8514 & 2017-1345) and registered with ClinicalTrial.gov (NCT#03539783).

### Subject Eligibility

Patients admitted to the PICU <18 years of age who were invasively mechanically ventilated and meeting consensus PARDS criteria ^1^ or who were admitted to the PICU without apparent lung disease and with expected duration of hospital admission 7 days or more were eligible for enrollment. PARDS is classified as mild, moderate or severe based on oxygenation impairment. We used oxygenation index for subjects with and oxygenation saturation index for subjects without an arterial line to quantify oxygenation impairment ^21^. Exclusion criteria were orders for limitation of resuscitation, known nasal pathology, a high risk of nasal bleeding as determined by the clinical team, or baseline oxygen requirement of 2 liters per minute or more. Enrollment was permitted at any time during PARDS course with earlier enrollment encouraged.

Inferior turbinate brushings and serum samples were collected on study days 1, 3, 7, and 14. Clinical and biometric parameters at the times of enrollment and specimen collection were recorded in a RedCap database.

### Standard Library Preparation and Sequencing

Specimen ribosomal RNA was depleted using RiboZero (Illumina) and libraries created using NexteraXT (Illumina), and sequenced on a HiSeq2500 using paired end sequencing of 150 base pairs at a sequencing depth of 10 million reads per sample.

### Unsuccessful RNA Amplification

Specimen ribosomal RNA was depleted using NEBNext rRNA Depletion kit (New England Biolabs) and amplified using SeqPlex (Sigma) with 20 cycles of amplification. Barcoding was performed first with PlexWell96 (SeqWell) and then with NexteraXT. Sequencing was not undertaken due to low cDNA concentration.

### Low-Input Library Preparation

Specimen RNA was amplified and barcoded using the NEBNext SingleCell/Low Input RNA Library Prep kit (New England Biolabs) per manufacturer instructions with 20 cycles of amplification using NEBNext multiplex oligos for barcoding. Specimen DNA concentration was normalized, and sequencing was performed using a Novaseq sequencer and a single S4 flow well with paired end sequencing of 150 base pairs each yielding ~ 10 million reads per sample.

### Statistical Analysis

Clusters were defined using Euclidean distance and principal component analysis in DESeq2 version 1.22.2. ^22^ For upstream analyses, ToppGene ^23^ and Ingenuity Pathway Analysis (v 20.0)^24^ were used with Bonferroni-corrected p-value (q-value) of less than 0.1 considered significant. R-v3.5.3 was used for all statistical analyses using ggplot2 and finalfit v-1.0.0 ^25^ with p-values of <0.05 considered significant using Fisher Exact test for nominal and ordinal data and Wilcoxon-Rank Sum and Kruskal-Wallis tests with Dunn’s *post hoc* text for continuous data.

## 5 RESULTS AND DISCUSSION

### Recruitment and Specimen Collection

From January 1, 2018 to November 30, 2019 we enrolled 26 control and 25 PARDS subjects. From these subjects, we collected 111 nasal brushings. PARDS subjects differed from control in illness severity (PELOD2), severity of lung injury, length of PICU and hospital stay, and exposure to corticosteroids (Table 1).

### Reasons for Failure of Non-low Input RNA Techniques

We arrived at our current protocol after several failures which are described in the supplement. Library creation was successful in only 13 of 45 (29%) using standard techniques and in 95% (61/64) of specimens using a low-input (NEB) approach. The sequences from these specimens was used for downstream analysis (Figure 1).

**Figure 1:**
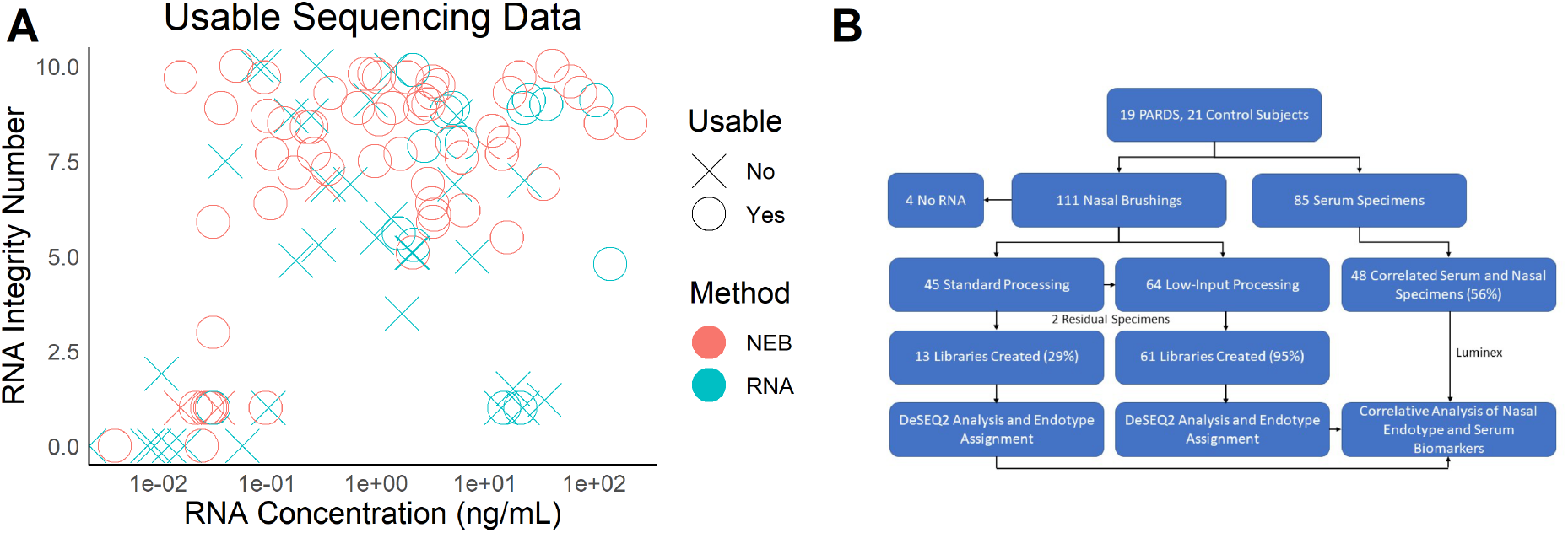
Study Specimens. (A) Comparison of standard (RNA) and low-input (NEB) techniques of nasal brushing cDNA library creation. NEB permitted creation of libraries from specimens of lower RNA concentration and lower quality (RNA Integrity Number). (B) Study specimen flow diagram.

### Assessing Nasal Specimen Similarity

Sixty-four nasal brushing specimens from 15 PARDS and 10 control subjects collected on study days 1, 3, 7, and 14, were analyzed by DESeq2. On days 1 and 3, four clusters were readily apparent by principal component analysis with two of these clusters (A1 and A2) containing control subjects (Figure 2 A&B). On study days 7 and 14, fewer samples were available for analysis, but clusters A1, A2, and C were still apparent (Figure 2 C&D). Analysis of all specimens together yielded the same specimens in clusters B and C, but specimens from A1 and A2 were no longer distinct and were combined for downstream analysis (cluster A, Figure 2 E). By Euclidean distance of the combined specimens, cluster B was clearly distinct from A and C, and the specimens in cluster C were grouped together, but cluster C was located within the larger cluster A (Figure 2 F). PARDS severity is classified as mild, moderate, or severe based on impairment in oxygenation^1^. While cluster A contained several specimens obtained during moderate or severe PARDS, clusters B and C were exclusively composed of such specimens (Figure 2 G). By linking the temporally obtained specimens for each subject, a clear trajectory from cluster B to cluster A and from cluster C to cluster A (with one exception) was noted (Figure 2 H). Lastly, we analyzed the 13 specimens collected and processed without amplification and with standard cDNA library prep using the same analysis method. Three groups were again identified, although not all control specimens were clustered together (Figure 3). Notably however, among the two control subjects in cluster C one developed PARDS, and one nearly met mild PARDS criteria (oxygen saturation index 4.8). Our initial practice of only enrolling invasively mechanically ventilated control subjects likely contributed to this finding. This longitudinal data with validation in a second dataset indicate that nasal transcriptomic gene expression in PARDS patients can be classified into one of three gene expression profiles. One similar to that of control subjects (Group A), and two others that are largely distinct from those of control subjects (Groups B&C).

**Figure 2:**
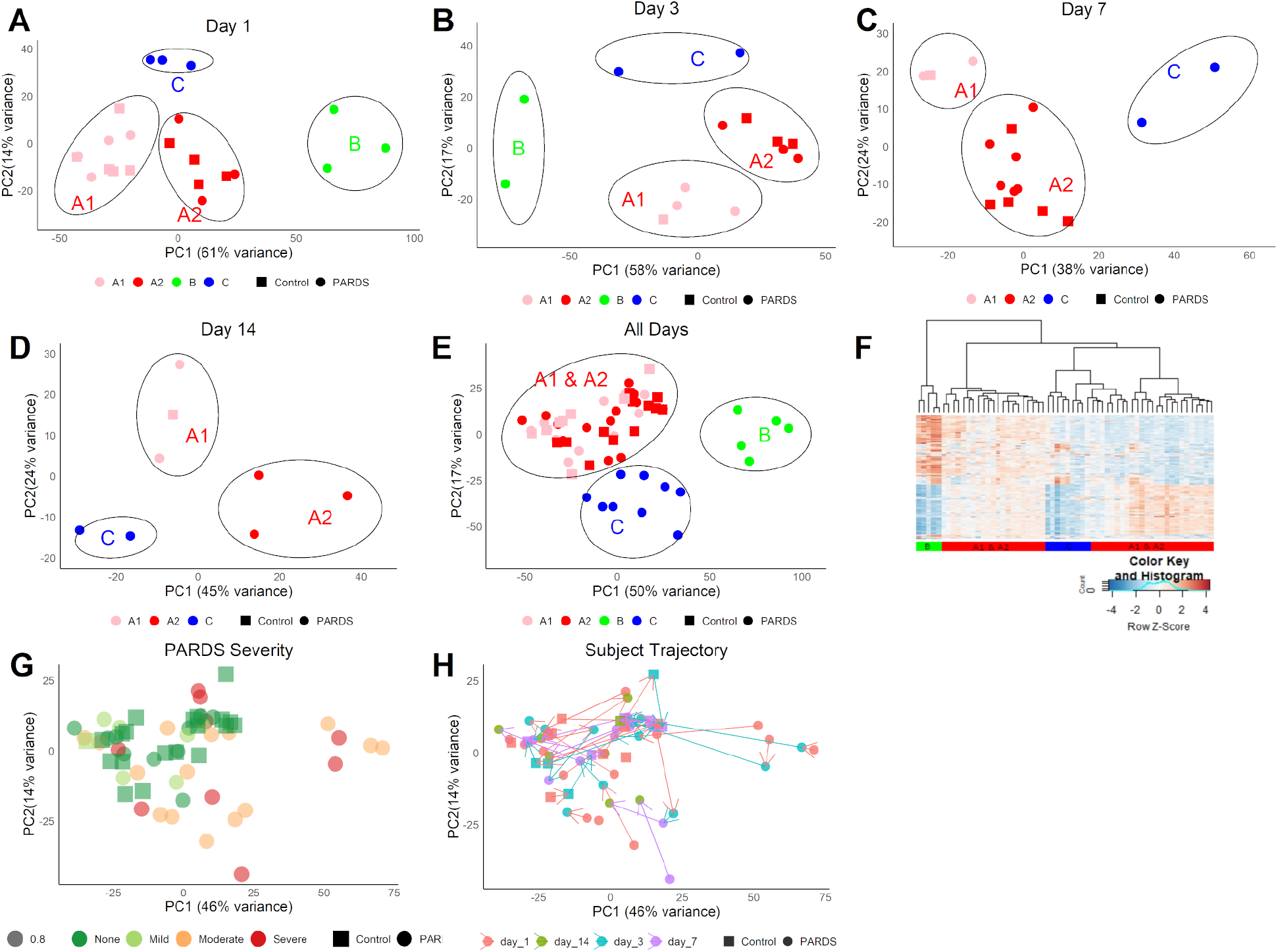
Nasal Specimen Transcriptomic Cluster Analysis. (A) Principal component analysis (PCA) of mRNA-Seq data from day 1 and (B) day 3 nasal specimens showed four clusters, A1, A2, B, and C. Specimens from control subjects were restricted to clusters A1 and A2. (C) Day 7 and (D) day 14 specimens had only A1, A2, and C clusters. (E) In the combined dataset, clusters A1 and A2 were no longer distinct but clusters B and C remained so. (F) A heatmap of Euclidean distance clearly differentiated the five PARDS specimens comprising cluster B. Specimens from cluster C were most similar to each other although not entirely separated from cluster A. A1 and A2 specimens were interspersed within A. (G) PARDS severity was greatest in specimens from clusters B and C although moderate and severe PARDS were present in cluster A. (H) Following specimens from each subject over time, subjects with specimens that were initially in clusters B and C moved into cluster A over time.

**Figure 3:**
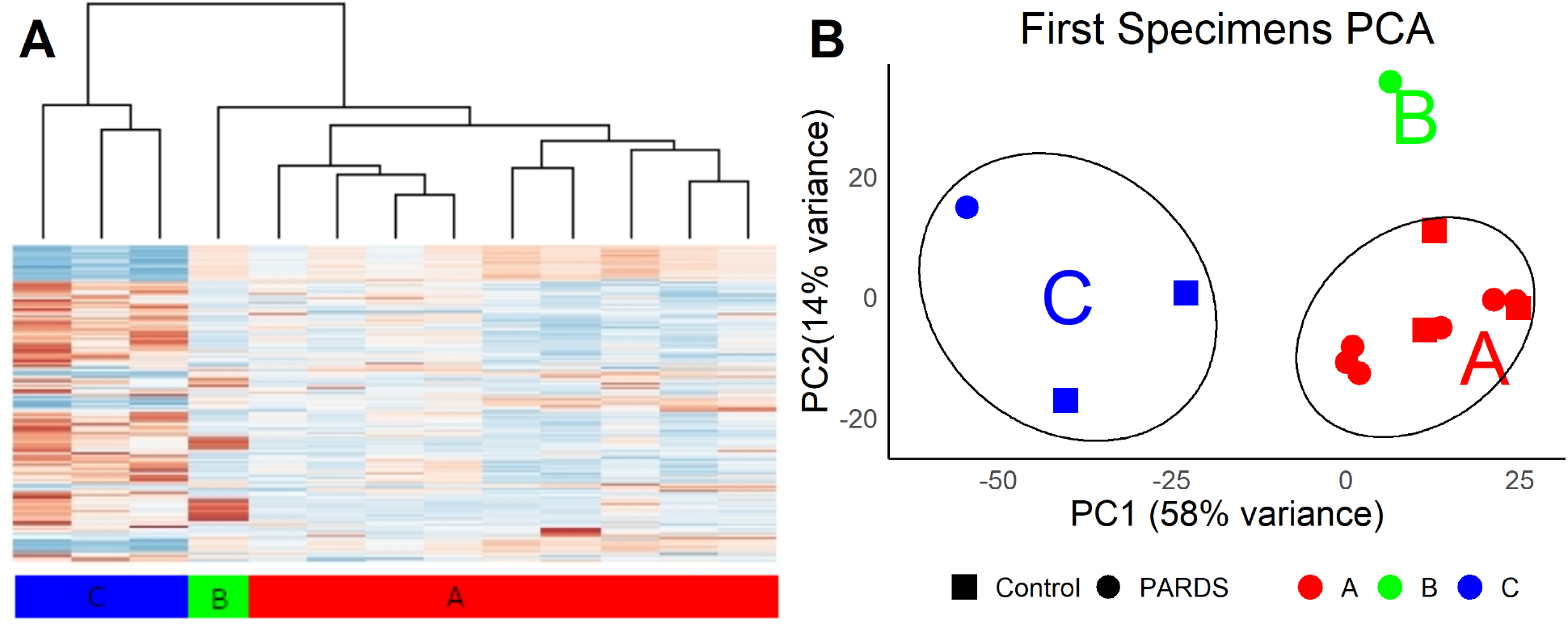
Similar Clustering in Distinct Group of Nasal Specimens. (A) Heatmap of the RNA processed by standard techniques for mRNA-seq (only 25% successful) showed clustering into three groups. (B) Similar to the low-input technique, specimens processed by this technique clustered into three groups. One control subject in group C developed PARDS, and the other nearly met PARDS criteria.

### Characterization of Three PARDS Endotypes

For this study, we define “endotype” as a biologically distinct disease subgroup that is clinically indistinguishable from other subgroups of that disease. To first determine whether or not there were clinical characteristics of the subjects or clinical measures present at the time of specimen collection that were predictive of cluster assignment, we performed a descriptive analysis of subjects (assigning subjects as “B” or “C” if any one specimen was so classified and “A” if all specimens were cluster A) and of specimens. We then performed univariate analysis of group A vs combined Group B&C, and multivariate logistic regression for any variables with p<0.1. For categorical variables Fisher’s exact test was used for statistical comparisons and for continuous and ordinal variables Wilcoxon-Rank Sum test was used. In a descriptive comparison between Group A, B, and C subjects, only disease severity (PELOD2) was statistically significant (Supplemental Table 1), and for individual specimens, PARDS classification (None, Mild, Moderate, or Severe), the presence of direct lung injury, and a viral or combined viral/bacterial cause of ARDS were significantly different between groups A, B, and C (Supplemental Table 2). In subject univariate analysis, group assignment (Control or PARDS) neared statistical significance (p=0.06) and both PELOD2 and PARDS severity were significantly higher in group B&C compared to group A (Supplemntal Table 3). In specimen univariate analysis, PELOD2 and direct lung injury neared statistical significance (both p=0.05), and PARDS severity, viral infection, and combined viral/bacterial infection were significant (Supplemental Table 4). For multivariate analysis by subject, we analyzed variables with p<0.1, and no clinical variable was significantly associated with Group A or the combined Group B&C (Supplemental Table 5). In multivariate analysis of specimens, PARDS severity (p=0.005) and viral PARDS trigger (p=0.04) were significantly associated with Endotype B or C (Supplemental Table 6). Although these findings (Figure 4) should be interpreted with caution due to low numbers, these data demonstrate that (1) none of the collected demographic or disease-specific data apart from a viral PARDS trigger significantly influences group assignment and (2) changes in the nasal transcriptome mirror changes in disease severity in PARDS subjects. Notably, an equal percentage (67%) of specimens classified as group B or C had a viral ARDS trigger. Although it did not meet statistical significance thresholds, mortality in Groups B&C was 27% compared to 5% in Group A (p=0.12 in univariate analysis) suggesting a clinically meaningful difference between the groups.

**Figure 4:**
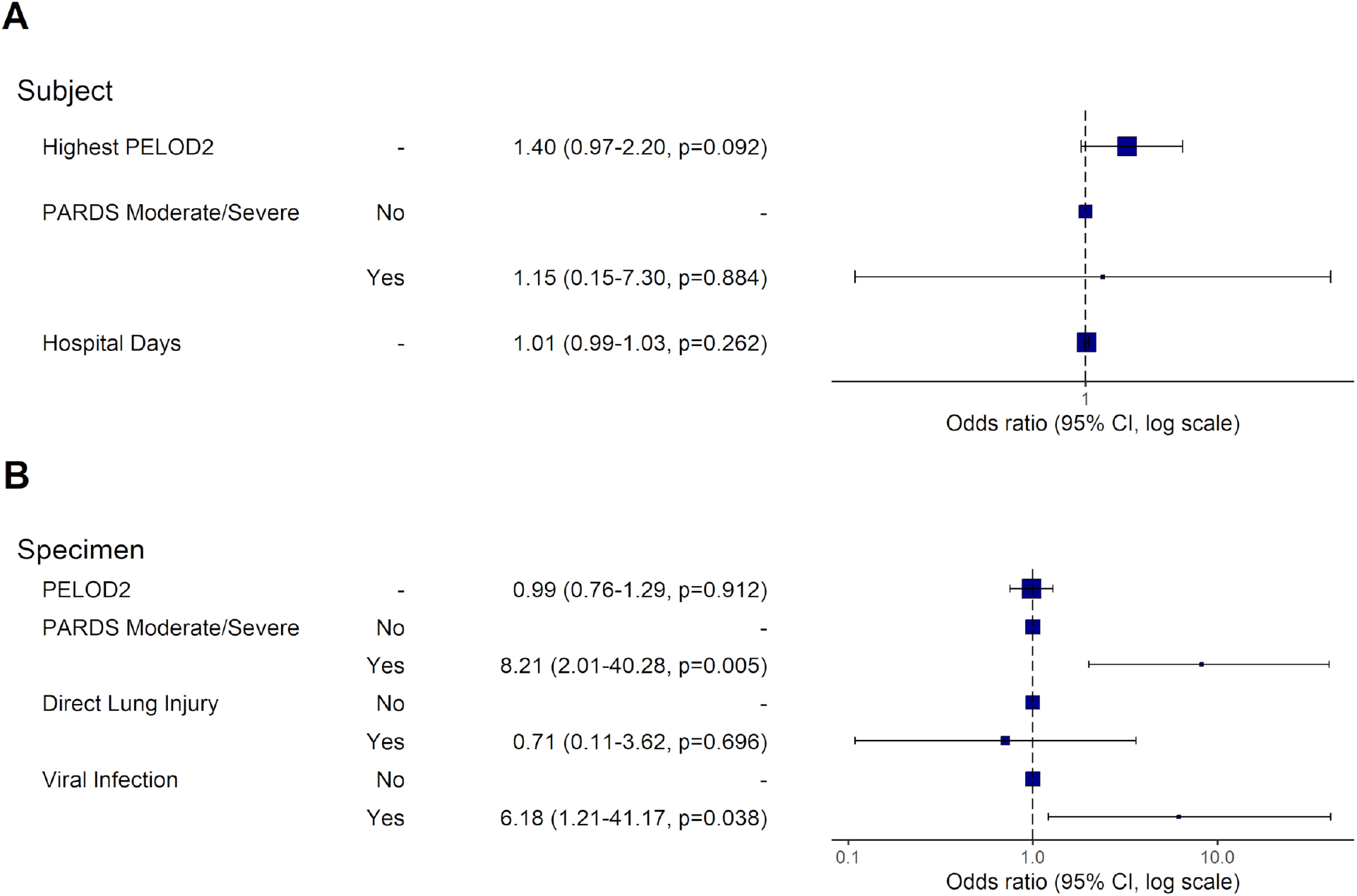
Variables Associated with Group B or C in Multivariate Logistic Regression. (A) Variables with p<0.1 in univariate analysis were subjected to multivariate analysis. Subjects with any specimens classified as B or C tended to have greater organ dysfunction, but this finding was not statistically significant. (B) Specimens from subjects with moderate or severe PARDs or with viral infection as a PARDS trigger were more likely to be classified as group B or C.

Since specimens from groups B and C were collected from subjects at times of greater lung injury severity and group A contained the specimens from these same subjects at times of lesser lung injury severity in addition to control subjects, we analyzed sets of deferentially expressed genes (DEGs) of group B compared to group A and group C compared to group A without respect to collection day. After initial filtering for expression level and identification of genes with adjusted p-value less than 0.1 with a fold-change of 2 or greater, the Group B analysis contained 1192 up and 873 downregulated DEGs. Group C contained 533 up and 86 downregulated DEGs. Toppgene analysis of these datasets revealed that compared to Group A, Group B had decreased and Group C had increased representation of genes related to microtuble dynamics and ciliary function. Group B had increases in many genes related to innate immunity (Figure 5 A-B). The pathways and gene family overepresented in the two datasets contained terms related to neutrophil degranulation, cytokines, and tumor necrosis factor for Group B and cilium assembly and dyneins for Group C (Figure 5 C-D). There was a notable absence of adaptive immune terms in this ToppGene analysis. Ingenuity Pathway Analysis was undertaken to identify potential regulators. Interferon-*γ* and tumor necrosis factor-related signaling were notable in Group B (Figure 5 E, G), and in analyzing network depth, suppression of FOXP1 and the proteosomal protein UBE3C were identified as potential master regulators of the Group B inflammatory cascade (Figure 5 F). There were no strong upstream regulators identified in Group C. Considering the differences in nasal epithelial cell gene expression in groups A, B, and C in conjunction with our findings that the only clinical variables that differentiated specimens from groups B and C from A were severity of lung injury and a viral cause of ARDS, and that viruses were the most common trigger of ARDS in both groups B and C, these data demonstrate that nasal epithelial transcriptomics can identify three distinct endotypes in PARDS: Endotypes A, B, and C.

**Figure 5:**
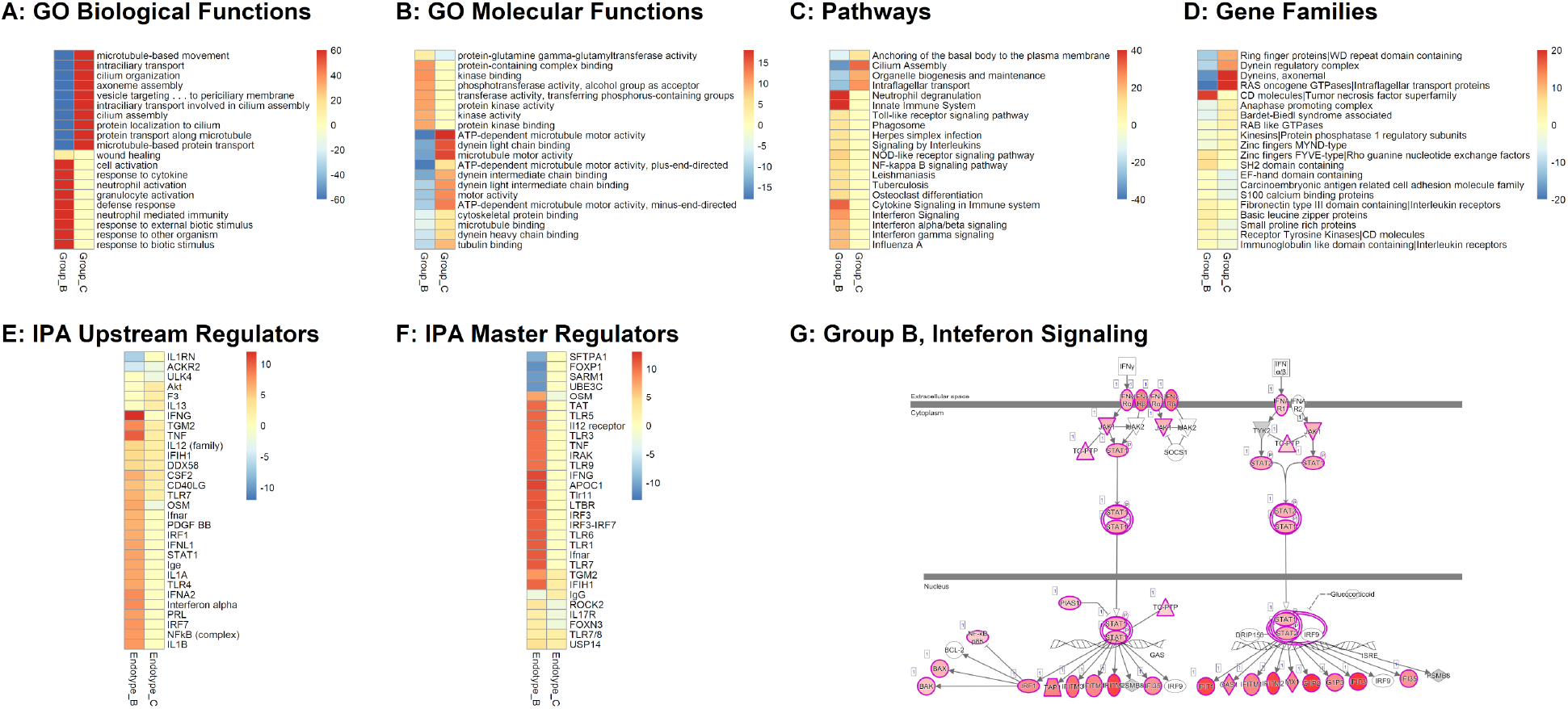
Characterization of ARDS Endotypes. (A) ToppGene and Ingenuity Pathway Analysis (IPA) were used to analyze DEGs in groups B and C compared to group A, which contained all of the control subjects in the dataset analyzed (the original dataset could not be normalized to the larger, second dataset because of differences in RNA processing and amplification). Innate immune processes were increased in group B and microtuble-related processes increased in group C and decreased in group C. (B) Gene Ontogeny (GO) molecular function analysis demonstrated a similar up- and downregulation of dynein and microtubule motor processes in groups C and B respectively. (C) Pathway and (D) Gene Family analyses were consistent with an increase in innate immune processes in group B and cilia-related ones in group C. All color scales for panels A-D represent -log10(Bonferroni-corrected p-value) for upregulated and log10(corrected p-value)for downregulated terms. (E) IPA analysis of upstream regulators showed increased Interferon-*γ* and tumor necrosis factor in group B and increased Akt and Interleukin-13 signaling in group C. (F) Network depth analysis of master regulators identified a potential role for FOXP1 suppression in Group B in addition to upregulation of many members of inflammatory signaling cascades. Oncostatin-M (OSM) was the only term with reciprocal scoring between the groups and is known to regulate endothelial cell cytokine production. Color scales for panels E-F represent activation or inhibition z-score. (G) Upregulation of many components of the Interferon-*γ* signaling cascade is shown. Red represents degree of upregulation. Green is absent but represents downregulation.

### Possible Sub-endotypes in B but not C

We were interested in the lack of an adaptive immune signature in Endotype B. In analyzing Endotype B and C subjects separately, we found that subjects in Endotype B clustered together tightly with differences driven by adaptive immune genes (Figure 6 A), while in Endotype C there was no such clustering apparent (Figure 6 B). While preliminary, these data suggest that within the inflammatory Endotype B, there may exists sub-endotypes that may represent distinct inflammatory processes in these subjects. It seems unlikely that the sub-endotypes represent resolution of inflammation because the subsequent specimen for each subject in Endotype B was Endotype A.

**Figure 6:**
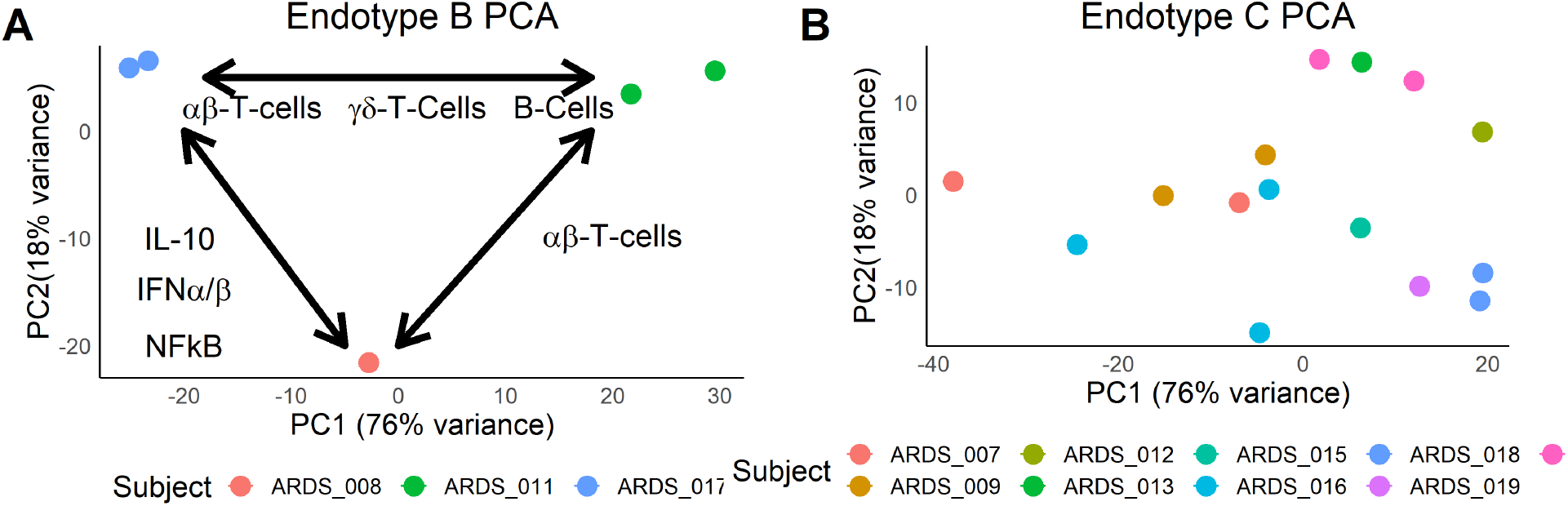
Sub-Endotypes in PARDS. (A) When analyzed separately, Endotype B specimens clustered by subject with pathways and cell-specific processes related to adaptive immunity differentiating them. (B) Subject separation was not noted for specimens in Endotype C.

### Comparison of Nasal Transcriptomic Endotypes with Published Serum Biomarkers

In both adults ^3,9,14^ and pediatrics ^11,13^, serum biomarkers have been reported to correlate with ARDS endotypes. We quantified many of these serum proteins to determine associations with nasal transcriptomic endotypes. Forty-eight serum samples collected on the same day as nasal brushing were available from six control and seventeen ARDS subjects. Interleukin-17 (IL17), Angiopoietin-2 (ANG2), IL6, IL17, IL18, Intercellular Adhesion Molecule-1 (ICAM1), Tumor Necrosis Factor-*α* (TNFa), and IL10 levels positively correlated with each other. Only 3 specimens had measurable levels of Interferon-*α*. Interestingly, several of these markers were anti-correlated with the alveolar type I cell marker Receptor for Advanced Glycation Products (RAGE) which was correlated with the neutrophil markers matrix metalloproteinase-9 (MMP9) and Myeloperoxidase (MPO). MMP9 and MPO did not correlate with the other neutrophil markers Granzyme B (GrB) and ICAM1 (Figure 7 A). Evaluating each of these biomarkers by group, GrB, ICAM1, Interferon-γ (IFNg), IL18, and Surfactant Protein D (SPD) were significantly elevated in ARDS but with many ARDS specimens having non-elevated levels for each (Figure S1). There was no significant correlation of any biomarker with PARDS severity (Figure S1). While only four subjects in Endotype B had serum samples available for analysis, TNFa was higher these specimens compared to specimens in Endotype A, and SPD was higher in Endotype C compared to Endotype A (Figure 7 B-G). These data are consistent with the concept that ARDS and PARDS represent a common clinical manifestation of different pathophysiological processes and that nasal transcriptomics can discriminate between them. Although the number of specimens is low, Endotype B had no clear association with other inflammatory biomarkers in the serum (Figure S1) as we hypothesized indicating that the inflammatory state of the nasal epithelium (and presumably the lower airway epithelium) is not necessarily correlated with the systemic inflammatory state. Elevated serum SPD levels have been associated with PARDS ^26^ and adult ARDS ^27–29^ severity. The higher level of SPD in Endotype C in conjunction with upregulation of processes related to microtubule function suggests that loss of barrier function is a characteristic of Endotype C.

**Figure 7:**
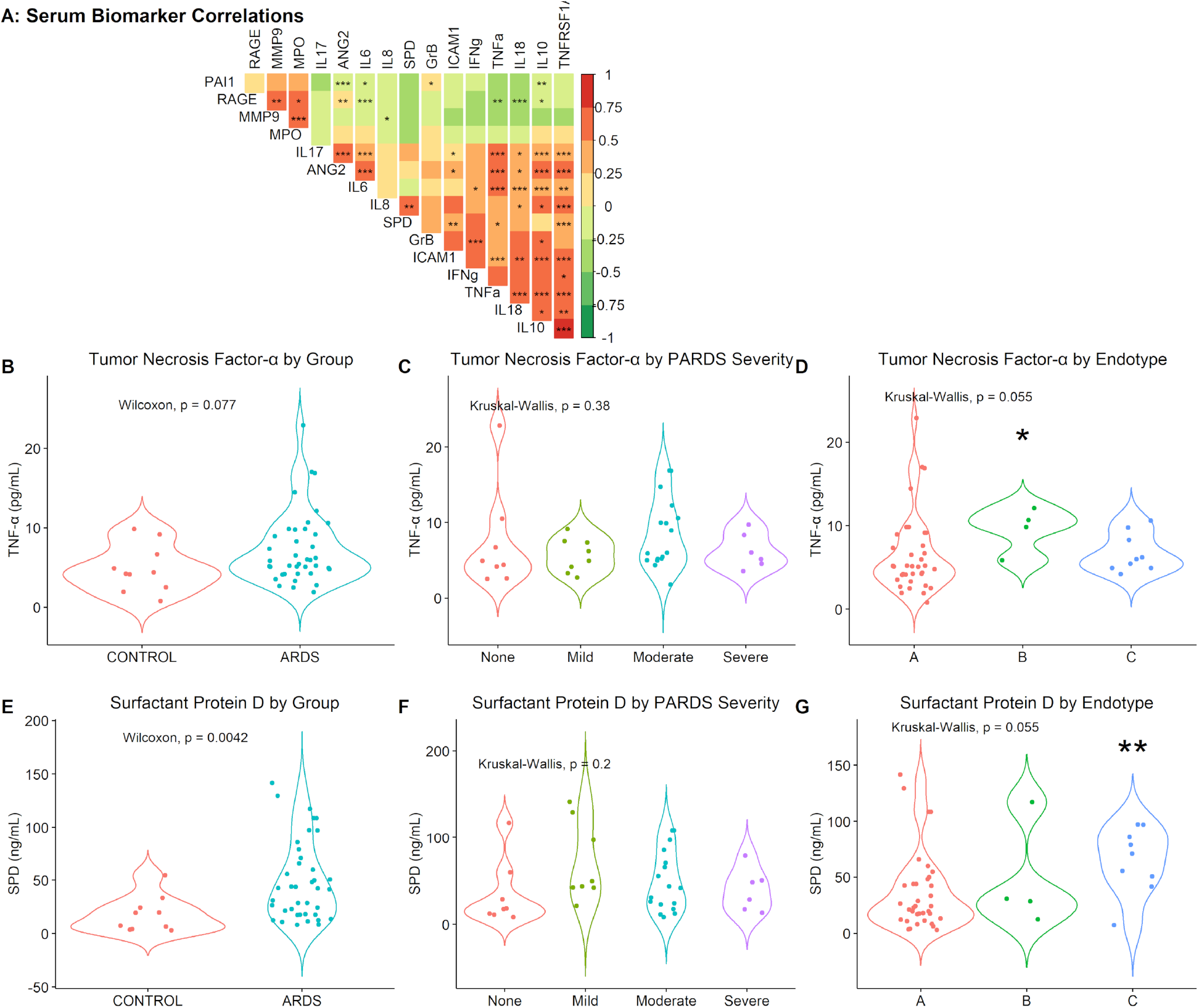
Serum Biomarkers and Nasal Transcriptomic Endotypes. (A) Forty-eight specimens from 23 subjects were evaluated for 16 biomarkers reported to identify ARDS and PARDS endotypes and Spearman-rank correlation coefficients were calculated for each as represented by color scale. * p<0.05, ** p<0.01, *** p<0.001 by Spearman Rank Correlation. (B) Tumor Necrosis Factor-*α* (TNFa) was in some PARDS subjects but (C) did not correlate with PARDS severity. (D) TNFa was higher in serum from Endotype B subjects. (E) Surfactant Protein D (SPD) was significantly increased in the serum of some PARDS subjects, but (C) this increase did not correlate with PARDS severity. (D) SPD was significantly higher in Endotype C compared to Endotype A. ** p<0.01 by Dunn’s *post hoc* test. ANG2 = Angiopoietin-2, GrB = Granzyme B, ICAM1 = Intercellular Adhesion Molecule-1, INFg= Interferon-*γ*, IL=Interleukin, MMP=Matrix Metaloproteinase, MPO=Myeloperoxidase, PAI1=Plasminogen Activator Inhibitor-1 (Serpin E1), RAGE=Receptor for Advanced Glycation End Products, TNFRSF1A=TNF Receptor Soluble Factor 1A.

## 6 CONCULSIONS

Our findings in conjunction with reports on the ability of serum biomarkers in pediatric ^11^ and adult ^9,14^ ARDS to identify clinically meaningful endotypes provide hope that future trials in ARDS can incorporate prognostic and predictive enrichment in subject recruitment and enrollment thus increasing the likelihood of discovering meaningful therapies. The lack of such enrichment could explain the frequency with which small pilot clinical trials show positive results only to fail in larger trials. Given sufficient infrastructure and support, nasal transcriptomics based endotyping could be performed in 48-72 hours to provide this diagnostic and prognostic enrichment.

Our findings of an inflammatory endotype (Endotype B) is in general agreement with the findings of Calfee, *et al*. ^14^ showing that hyper-inflammatory adult ARDS responds less well to high levels of PEEP and liberal fluid administration. However, except for TNFa, inflammatory serum biomarkers were not significantly elevated. Our findings suggest that the nasal epithelium harbors cell-specific information of adaptive immune processes that could be exploited for targeted immunotherapies. Endotype C was less distinct from Endotype A which contained control and resolved PARDS specimens, but the upregulation of microtubule associated processes and cilia-related genes in this endotype paired with their downregulation in Endotype B is interesting. One hypothesis is that these changes reflect altered ciliary function. This possibility is intriguing because *β*-agonists improve ciliary function. It is perhaps this aspect of ARDS and not enhanced alveolar fluid resorption that drive the positive findings in the BALTI-1 trial but were not replicated in BALTI-2 ^30^. Both were clinical trials of *β*-agonists in ARDS based on the hypothesis that increased epithelial cell cyclic AMP levels would enhance alveolar fluid reabsorption. A second possibility is that Endotype B has alterations in epithelial cell cytoskeletal function. Serum SPD levels are thought to be reflective of lung epithelial barrier integrity, and thus, it’s possible that Endotype C reflects a reduced barrier integrity phenotype. However, the reciprocal nature of these pathways in Endotype B raises the possibility that Endotype C represents recovery from Endotype B. We think this is unlikely because we observed one subject moving from Endotype A into C with worsening PARDS, we saw no subjects moving from Endotype B to Endotype C over time, and the length of time subjects remained in Endotype C was longer than Endotype B. Further characterization of these microtubule and ciliary findings in Endotype C is required.

Several study strengths and weaknesses are noted. First, additional subjects and specimens would increase the confidence that only three endotypes exist. Endotype B consisted of only four subjects. However, the identification of these same endotypes at different time points and in a largely distinct cohort of subjects and specimens increases our confidence that Endotypes A, B, and C represent true biologically distinct entities. Future studies should use a single early specimen from each subject to confirm these endotypes. Second, we did not restrict the time frame in which subjects could be enrolled. Our rationale for such an approach was that the underlying pathobiology of intubated ARDS subjects is well-established by the time of endotracheal intubation. However, it also likely introduced a greater number of subjects with resolving PARDS and caused an over representation of Endotype A. Third, we did not exclude subjects with baseline ventilator support or immunodeficiency so long as baseline oxygen requirement was less than 2 liters per minute. These patients are likely predisposed to worse PARDS with a given insult and this too may have caused over-representation of Endotype A. Lastly, although nasal transcriptomics reflect lung processes in other diseases, future studies should correlate nasal, bronchial, and bronchoalveolar lavage gene expression. A notable strength of our study that differentiates it from other ARDS endotyping studies to date is the inclusion of a critically-ill control group. This demonstrated that not all PARDS subjects have nasal gene expression profiles different from that of controls and that with PARDS resolution, the nasal transcriptome returns to normal.

## Data Availability

Once peer-review is completed and the publication is accepted, de-identified data will be made available to interested investigators.

## Acknowledgements

We would like to thank patients and their families for their participation in this study and nurses and respiratory therapists in the Cincinnati Children’s Hospital Pediatric ICU for assistance with sample collection.

**List of abbreviations**
ARDS: Acute Respiratory Distress Syndrome
COPD: Chronic Obstructive Pulmonary Disease
ECMO: Extracorporeal Membranous Oxygenation
FiO2: Fraction of Inspired Oxygen
MAP: Mean Airway Pressure
OI: Oxygenation Index (FiO2 X MAP / PaO2)
OSI: Oxygen Saturation Index (FiO2 X MAP / SaO2)
PaO2: Arterial Partial Pressure of Oxygen
PICU: Pediatric intensive care unity
PARDS: Pediatric Acute Respiratory Distress Syndrome
RNA: Ribonucleic Acid
SaO2: Arterial Oxyhemoglobin Saturation

